# Willingness and acceptability of anxiety and depression screening among women living with HIV in Ethiopia

**DOI:** 10.1101/2020.06.15.20131466

**Authors:** Abdilahi Yousuf, Ramli Musa, Muhammad Lokman Md. Isa, Siti Roshaidai Mohd Arifin

**Affiliations:** College of medicine and health sciences, Jijiga University, Jijiga, Ethiopia; Department of psychiatric, Kulliyah of medicine, IIUM, Kuantan, Malaysia; Department of Basic Medical Sciences, Kulliyah of Nursing, IIUM, Kuantan, Malaysia; Academic and industrial linkages, Kulliyah of Nursing, IIUM, Kuantan, Malaysia

**Keywords:** Depression, HIV, AIDS, acceptability, screening

## Abstract

**Introduction:** Integration of related health services, such as screening of depression in HIV care is crucial for improving the quality of care and enhancing the use of scarce resources in developing countries. However, commonly these interrelated health services are commonly provided separately and there are many missed opportunities. Little is known about the client-related non-implementation issues. This study aims at examining the acceptability of anxiety and depression screening among women living with HIV.

**Material and methods:** This was a facility based cross-sectional study which included women living with HIV attending two hospitals in Jijiga town, Ethiopia. The study participants were identified using systematic random sampling method. An exit interview was conducted with the use of a pretested questionnaire. The gathered data was analysed using SPSS version 23 and multivariate logistic regression model was used to determine factors associated with the factors associated with the acceptance of anxiety and depression screening.

**Results:** A total of 409 women participated in this study. Though, only 115 (28.1%) were aware about the existence of anxiety and depression screening, 357 (87.3%) accepted to receive the screening for anxiety and depression. Requisite of partner approval was the most common reason for not accepting the screening of anxiety and depression 21 (40.4%). When used multivariate logistic regression model; holding college level education, divorced and were living without partner, being self employed by occupation, aware of the existing screening service, source of information from health care providers and history of previous screening were significantly associated with acceptance of depression screening.

**Conclusion:** This research has shown that women participant living with HIV were willing to undergo the screening for depression, hence future interventions should focus on the integration of mental health screening in HIV clinical setup.

## Introduction

Depression, a common mental disorder which affects more than 300 million individuals of all age groups in this world wide. This disorder which is more common in women than men, is the primary cause of disability and one of the main contributors of global disease burden (1) including suicide (2,3).

Depression affects about 20% of women during their lifetime, with reproductive health problems being a period of high vulnerability (4). It is also predicted that depression which is estimated to be the second leading cause of disease and disability in the world in 2020, will be twice common in women than men (5–9).

Anxiety and depression is common among people living with HIV infection (5,10–18). High and low-income countries have presented higher rates of depression in people living with HIV when compared with HIV negative individuals. The level of depression mostly appears to be related to the severity of symptoms of HIV infection (19).

In developing countries, depression is also one of the main challenges that should be addressed when dealing with HIV prevention strategies, as the two illnesses are intensely associated and aggravate each other. In recent evidences, depression and depressive symptoms are found to be the key predictors of poor adherence to HIV treatment (20) and negatively impact on clinical outcomes (21).

So far, depression is highly prevalent in Ethiopia, and it is considered as one of the common psychiatric disorders affecting women as its associates are gender-specific (5–10,14,22,23). One of the findings conducted in Ethiopia showed that there is high prevalence of depression on people living with HIV/AIDS and stated that 38.94% of the studied participants were depressed (7). Another systematic review conducted in Ethiopia has presented that the pooled prevalence of depression was higher among People Living with HIV than the general population (24).

Regardless of its prevalence among people living with HIV, depression and anxiety are commonly under diagnosed by health care providers. More than half of those eligible patients, miss the opportunity to be screened for anxiety and depression disorders by doctors (25).

Furthermore, people with chronic illnesses are usually unwilling to seek health professional support and counselling. A mong people who manifest depressive symptoms, less number is reported to seek treatment in their course of disorder (26,27).

Even though the significance of depression screening for the intervention and care of HIV patients in health facilities has been long recognized, yet little is known about the acceptability and predictive factors related with the acceptance of anxiety and depression screening among women attending HIV care services. Therefore, this study will examine the acceptability and the predictive factors associated with screening of anxiety and depression among women Living with HIV. The findings of this study can be a baseline for the introduction of anxiety and depression screening in HIV care services of health facilities in Ethiopia.

## Methods and materials

### Study design

A facility-based cross-sectional study was carried out in Antiretroviral Therapy clinics of two hospitals. This is study was the first phase of extensive mixed-method research conducted among women with HIV.

### Study population

The study population was women aged 18 years or older who are attending HIV treatment service in two hospitals.

### Sample size determinations and sampling procedure

A single population proportion formula, was used to estimate the sample size of women participants to be studied with a 95% CI and 5% margin of error. (P) the prevalence of depression from a baseline study in Addis Ababa, Ethiopia (i.e. 41.2% of studied respondents were depressive). Expecting a 10% of non-response rate the total sample size was 409 patients.

A systematic sampling procedure was used for the selection of the study participants. To determine the sampling interval, the total study population who had an appointment during the data collection was divided by the total sample size, then the initial point was randomly selected.

### Data collection

A trained nurses conducted an interview with women attending the antiretroviral treatment centres. A pretested structured questionnaire was used. Participants were asked about their socio-demographic status, their clinical and laboratory investigation results were also retrieved and questions regarding about their awareness on depression screening, their previous test and their acceptance of the screening were asked.

### Screening

For the initial screening of the patients, a Hospital Anxiety and Depression Scale (HADS) was used. It is a tool developed my Zigmond and Snaith to be practiced for general population. Unlike other depression screening tools, this questionnaire is easy to deal separately any comorbidity or impairment secondary to another disease (28). This assessment tool has seven items for anxiety and seven items for depression and it takes mostly to finish around 2 to 5 minutes. As recommended, both anxiety and depression separately were scored. For the scales of anxiety and depression; scores which are less than 7 shows that the participants are not depressed, for the score 8-10 are mild cases and 11-14 is reported as moderate and 15-21 scores shows severe cases. This questionnaire is validated and published in deferent languages including Ethiopian local language.

### Data analysis

The gathered data was analysed using IBM SPSS statistics version 22. A univariate analysis was conducted using chi-square statistics to identify the predicting factors associated with the outcome variable, acceptance of anxiety and depression screening. Multivariate logistic regression was performed to identify independent variables associated with the outcome variable. Variables with p-value 0.005 or less were included in the model. Odds Ratio (OR) and Adjusted Odds Ratio (AOR) with their Confidence Interval (95%CI) were calculated to determine the strength of the association.

#### Ethical issues

Ethical approval of the study was obtained from International Islamic University of Malaysia Research Ethics Committee (IREC). Permission letter was also gained from Jijiga University board of review. A written informed consent was attained from each study participant after clarifying the purpose of the study & all other relevant information about the study. Privacy and confidentiality of women participants was strictly maintained.

## Results

### Characteristics of the respondents

A total of 409 women participated in this study. The mean age of the study participants was 34.5 years with standard deviation of 8.75 years. Majority of the respondents were urban dwellers 377 (92.2%) and Amhara by ethnicity 144 (35.2%). Among the total participants, 129 (31.5%) were married, 195 (47.7%) were followers of Orthodox Christian. Regarding the educational background, most of them 190 (46.5%) did not attend any formal education, almost half of the respondents 207 (50.6%) were unemployed, and almost half of them 206 (50.4%) reported their family income was less than 1400 Ethiopian birr which is equal to 37.5 US dollars. Among the interviewed participants, almost half of them were nulliparous women 208 (50.9%) as shown in (Table 1).

**Table 1.** Socio-demographic characteristics of participants (n= 409).

| Variable | Categories | Frequency | Percent (%) |
| --- | --- | --- | --- |
| **Residence** | Urban | 377 | 92.2 |
|  | Rural | 32 | 7.8 |
| **Age** | 15-24 | 57 | 13.9 |
|  | 25-49 | 339 | 82.9 |
|  | >49 | 13 | 3.2 |
| **Marital Status** | Married | 129 | 31.5 |
|  | Single | 120 | 29.3 |
|  | Widowed | 82 | 20.0 |
|  | Divorced | 78 | 19.1 |
| **No of living children** | Nulliparous/No children | 208 | 50.9 |
|  | 1-3 | 160 | 39.1 |
|  | 4-6 | 41 | 10.0 |
| **Education** | No formal education | 190 | 46.5 |
|  | Primary education | 97 | 23.7 |
|  | Secondary level | 83 | 20.3 |
|  | College/University | 39 | 9.5 |
| **Ethnicity** | Amhara | 144 | 35.2 |
|  | Somali | 137 | 33.5 |
|  | Oromo | 86 | 21.0 |
|  | Southern Nations | 42 | 10.3 |
| **Religion** | Orthodox | 195 | 47.7 |
|  | Islam | 165 | 40.3 |
|  | Catholic | 12 | 2.9 |
|  | Protestant | 37 | 9 |
| **Occupation** | Un employed | 206 | 50.4 |
|  | Self employed | 117 | 28.6 |
|  | Student | 44 | 10.8 |
|  | Govt employee | 42 | 10.3 |
| **Income in ETB** | <1400 | 206 | 50.4 |
|  | 1400-3800 | 129 | 31.5 |
|  | >3800 | 74 | 18.1 |

**Table 2:**
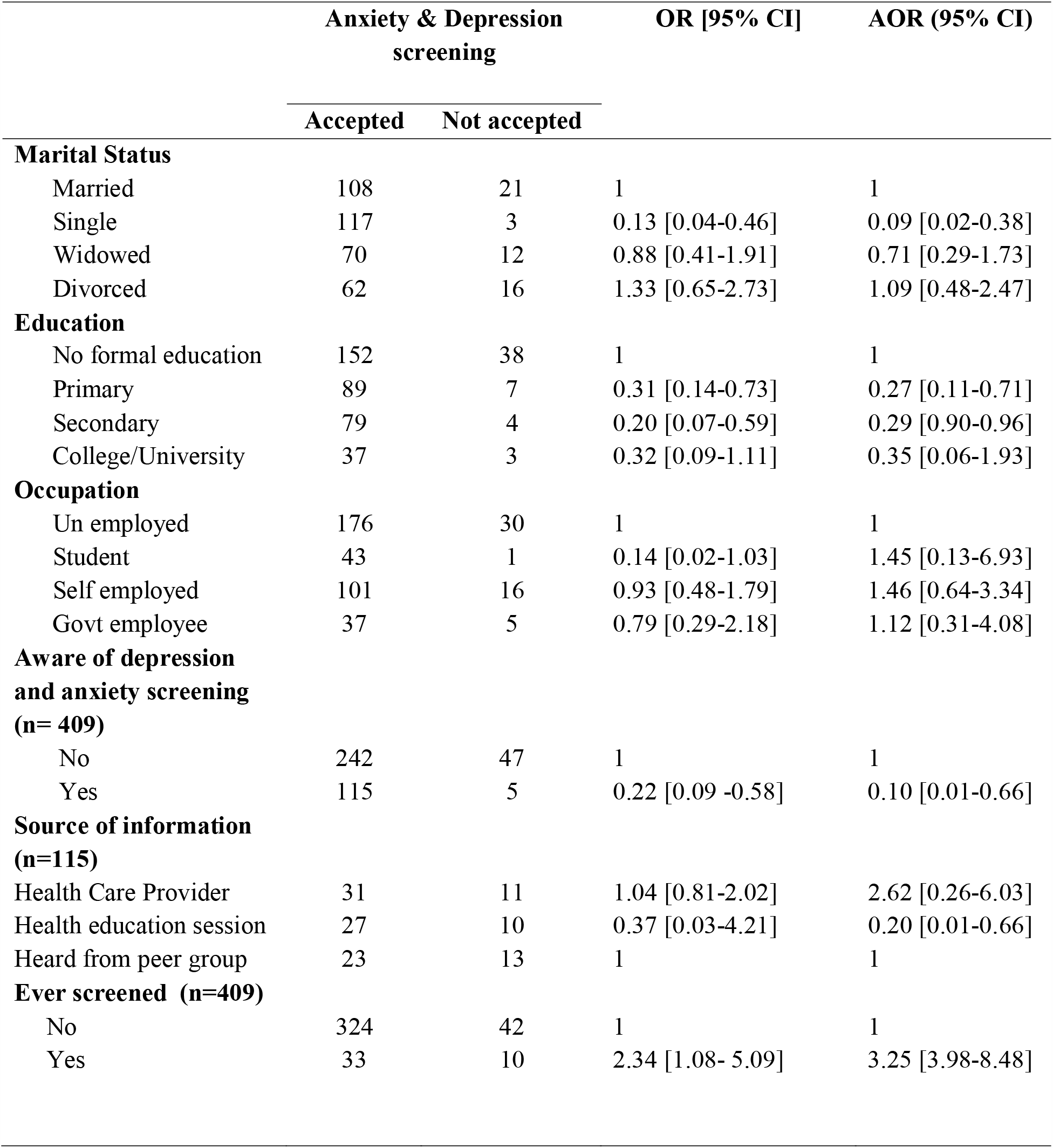
Association between socio-demographic and clinical characteristics with acceptance of depression and anxiety screening among participants.

|  | Anxiety & Depression<br>screening |  | OR [95% CI] | AOR (95% CI) |
| --- | --- | --- | --- | --- |
|  | Accepted | Not accepted |  |  |
| **Marital Status** |  |  |  |  |
| Married | 108 | 21 | 1 | 1 |
| Single | 117 | 3 | 0.13 [0.04-0.46] | 0.09 [0.02-0.38] |
| Widowed | 70 | 12 | 0.88 [0.41-1.91] | 0.71 [0.29-1.73] |
| Divorced | 62 | 16 | 1.33 [0.65-2.73] | 1.09 [0.48-2.47] |
| **Education** |  |  |  |  |
| No formal education | 152 | 38 | 1 | 1 |
| Primary | 89 | 7 | 0.31 [0.14-0.73] | 0.27 [0.11-0.71] |
| Secondary | 79 | 4 | 0.20 [0.07-0.59] | 0.29 [0.90-0.96] |
| College/University | 37 | 3 | 0.32 [0.09-1.11] | 0.35 [0.06-1.93] |
| **Occupation** |  |  |  |  |
| Un employed | 176 | 30 | 1 | 1 |
| Student | 43 | 1 | 0.14 [0.02-1.03] | 1.45 [0.13-6.93] |
| Self employed | 101 | 16 | 0.93 [0.48-1.79] | 1.46 [0.64-3.34] |
| Govt employee | 37 | 5 | 0.79 [0.29-2.18] | 1.12 [0.31-4.08] |
| **Aware of depression**<br>**and anxiety screening**<br>**(n= 409)** |  |  |  |  |
| No | 242 | 47 | 1 | 1 |
| Yes | 115 | 5 | 0.22 [0.09 -0.58] | 0.10 [0.01-0.66] |
| **Source of information**<br>**(n=115)** |  |  |  |  |
| Health Care Provider | 31 | 11 | 1.04 [0.81-2.02] | 2.62 [0.26-6.03] |
| Health education session | 27 | 10 | 0.37 [0.03-4.21] | 0.20 [0.01-0.66] |
| Heard from peer group | 23 | 13 | 1 | 1 |
| **Ever screened (n=409)** |  |  |  |  |
| No | 324 | 42 | 1 | 1 |
| Yes | 33 | 10 | 2.34 [1.08- 5.09] | 3.25 [3.98-8.48] |

### Acceptance of anxiety and depression screening

Among the 409 interviewed participants, only 120 (29.3%) were aware about the existence of anxiety and depression screening. Majority of them 50 (41.7%) mentioned that the HCP have introduced them screening. The other sources of information included health education sessions 32 (26.7%), followed by information obtained from other peer members 38 (31.7%). When asked about their experience of previous screening, most of the women 366 (89.5%) claimed that they had never been tested previously. However, majority of the participants, 357 (87.3%) accepted to receive the screening. The remaining respondents presented various reasons for their withdrawal of the screening which included requirement of partner approval (40.4%), longer waiting time (38.5%) and fear of stigma (21.2%).

### Factors associated with the acceptance of anxiety and depression screening

To identify the predicting factors of acceptance of depression and anxiety screening, we analysed the association between socio-demographic characteristics, awareness of the existing screening, history of previous screening and acceptance of women participants.

Majority of the participants who accepted the screening were holding college level education (OR: 0.32; 95 % C.I: 0.09-1.11), divorced (OR: 1.33; 95 % C.I: 0.65-2.73), were self employed by occupation (OR: 0.93; 95 % C.I: 0.48-1.79), aware of the existing screening service (OR: 0.22; 95 % C.I: 0.09-0.58), and were offered the screening by health care providers (OR: 1.04; 95 % C.I: 0.81-2.02). Moreover, a larger proportion of the participants who accepted the screening had a history of previous screening of depression and anxiety (OR: 2.34; 95 % C.I: 1.08-5.09) as shown in (Table: 2).

To adjust for potential cofounders, a multivariable logistic regression was used. Participants who had a college level of education were more likely to accept the screening offer as indicated in (Table: 2). Being aware about the screening service was also associated with accepting the screening and respondents that were invited to the screening by HCP were two times more likely to accept the test than those who heard from other source of information. In addition to this, women who had a previous history of screening practice were three times more likely to accept the screening.

## Discussion and conclusion

In this study we examined the acceptance of anxiety and depression screening among women living with HIV who attend the antiretroviral treatment care. The findings of this study has shown that the acceptance rate for screening of anxiety and depression among the study participants was 87.3% and this indicates relatively lower proportion when compared to a study conducted on depression and anxiety among pregnant women in Kilimanjaro, Tanzania in which 96.2% of the respondents accepted the screening. (11) and another similar study on prevalence of depressive symptoms on women in Sabah, Malaysia were 92.2% in which women accepted to undergo screening for the study (29).

Probably, the reason for the variation of the findings might be the difference in the study population, since the later studied on pregnant women who possibly had more frequent visit and follow up in their health facilities. One unanticipated finding shows regardless of the acceptability level of the participants, the awareness of the respondents about the availability of anxiety and depression screening was low (28.1%), when compared to other studies conducted earlier in different settings. The reason could be explained non-existence of proper mental health education sessions in the facility where the patients were expected to receive information.

The results of this study shows that the majority of the participants who accepted the screening were holding college level education. Hereafter, education denote the formal education obtained in academics and not merely the health related knowledge of the women participants. This finding is consistent with other studies which presented that positive association between higher educational level and women’s attitude towards anxiety and depression interventions (30,27). It is common in Ethiopia, that low service utilization is typically associated with the poor literacy among women. In line with this, it is found that HIV patients with no formal education was more likely to develop distress (31). As a result, education and awareness pay a major role in reducing the unmet needs in health service utilization including depression screening.

The respondents who were divorced, and those who were self employed by occupation were more likely to accept the screening. Contrary to expectations, and not common in low income countries where women empowerment is low, this finding may determine the autonomy of women for taking decisions in regard to health seeking when they are economically independent (32). However, among the married women, their claim of partner approval was the main reason of opting out the screening offer. Thus, in marital union, male involvement in decision making for the screening uptake was acknowledged.

Most of the study participants has mentioned health care provider’s as their main source of information to take part the screening. Although, this study did not initially identify the reasons for the high acceptance rate, the health provider’s initiated counselling could be the motive for the high acceptance rate. One of the HCP’s drive could be ascribed to their intention to practice the protocol of Ethiopia’s national guideline for comprehensive HIV prevention, care and treatment which suggests the screen for depression among the HIV patients in regard to. (33). Despite, the HCPs recognize that the screening was prominently required service, however it was not practiced in the studied health set up. This is common in major health facilities where depression is often unnoticed and not practiced (34). This could be attributed to the reality on the ground which is related with the practicality, availability of validated tools, less number of trained professionals, and overall the guiding principle of the hospital implementation plan. Depending on the regional difference, these limited service provision was recognized to increase a large proportion of patients, remain un-detected in the ART.

Moreover, a larger proportion of the participants who accepted the screening had a history of previous screening of depression and anxiety. This similar to other studies which concluded that women who had a previous engagement in screening were more receptive and willing to accept the screening (35–37). Yet, the type of screening tool and their decision process determines their choice to accept the screening. As several studies has published that women understandings and decision manners commonly regulates their response and acceptance of the screening (38–40). Thus, in order to reduce the frequent challenges of integrating mental health with the comprehensive HIV care and minimize the unmet need of HIV women to be screened for depression, recent findings suggest that ART attendees should be invited for depression screening and eligible clients should be linked for further examination and treatment (41).

## Conclusion

As the screening of symptomatic depression in HIV patients has recorded an apparent health implication for the adherence of treatment and care, health care providers should consider a routine check-up for the HIV patient’s neurocognitive functioning and wellbeing. This research has shown that women participant living with HIV were willing to undergo the screening for depression and anxiety. Therefore, all concerned bodies like health organizations should enhance efforts that promote mental health interventions in order to address vulnerable community member like people living with comorbid diseases; HIV and depression.

## Data Availability

The qualitative data included in this study is available from the authors upon request.

## Abbreviations

AIDS: Acquired Immune Deficiency Syndrome
ART: Antiretroviral Therapy
AOR: Adjusted Odds Ratio
HADS: Hospital Anxiety and Depression Scale
HCP: Health Care Provider
HIV: Human Immunodeficiency Virus
OR: Odds Ratio

## Ethics approval and consent to participate

All the study procedures were according the ethical standard of the institution’s human ethical procedure. The ethical approval of this study was obtained from the IIUM Research Ethics Committee (IREC) (Reference: IIUM/305/14/11/2/IREC/2018-042. Dated: 04/01/2019). A permission letter was also gained from the Jijiga University board of review. The women participants in this study were provided a written document approved by IREC and informed consent then confirmed by their signatures for the use and dissemination of the study findings.

## Consent for publication

Not applicable

## Data availability

The qualitative data included in this study is available from the authors upon request.

## Authors contribution

AY and SR: conception and design, data collection, statistical analysis, interpretation of data and drafting the manuscript. RM and ML: critical revision of the manuscript. All authors have contributed and approved this manuscript.

## Acknowledgements

The authors would like to acknowledge the WLHIV who participated this study and the medical staffs working in the ART for their assistance through the data collection process.

